# Wearable Gait Analysis is Highly Sensitive in Detection of Fatigue-Induced Exercise in Low Intensity Training Among University Football Players

**DOI:** 10.1101/2024.04.14.24305514

**Authors:** Seungmin Shin, Rakesh Tomar, Minji Son, Seoungeun Kim, Yongho Lee

**Affiliations:** Seowon university, South korea; King fahd university of petroleum & minerals, Saudi Arabia; JEIOS inc., South korea; Seoul national university, South korea

**Keywords:** Gait analysis, wearable sensor, IMU sensor, Fatigue

## Abstract

Gait analysis is crucial for understanding human movement patterns and detecting changes induced by factors such as fatigue. Fatigue can significantly impact gait dynamics, especially in athletes engaged in low-intensity training sessions like university football players. This study aimed to investigate the sensitivity of wearable sensors in detecting fatigue induced by low-intensity football training among university players. Twenty healthy male university football players participated in the study, undergoing gait analysis using wearable sensors before and after a 90-minute football training session. Data were collected using shoe-type IMU sensors for gait analysis, GPS trackers for exercise monitoring, and heart rate monitors for heart rate assessment. Participants also reported their perceived exertion using the Borg RPE scale. Results showed significant changes in various gait parameters post-exercise, including decreased cadence, increased percentage of double support, decreased percentage of single support, and increased time of toe-off. However, parameters like stride length remained unchanged. Center of gravity parameters did not show significant differences except for an increase in the ML(Y) acceleration post-exercise. The study suggests that even low to moderate-intensity exercise can induce fatigue, impacting walking dynamics. Wearable IMU sensors proved highly sensitive in detecting accumulated fatigue, even in low-intensity exercises, providing valuable insights into athletes’ physical deterioration during daily activities. This method could be crucial for monitoring fatigue and preventing injuries among athletes engaged in various sports activities. Further research is recommended to explore the impact of fatigue on other gait features and to evaluate gender differences. Additionally, examining muscle phosphocreatine readings could provide further insights into fatigue-related changes in gait. Nonetheless, the study highlights the effectiveness of wearable IMU sensor gait tests in detecting fatigue induced by low to moderate-intensity exercises, emphasizing the importance of monitoring fatigue for injury prevention and performance optimization in athletes.

## Introduction

Gait is a significantly coordinated movement. Gait involves raising the swinging leg, moving leg forward, and moving center of gravity (COG) from between the legs to the stance foot in order to generate the necessary forward momentum using the supporting leg [1]. During gait initiation, it is important to shift your weight between your legs and maintain balance as you shift your weight [2]. The rapid lateral weight transfer during swing leg lifting poses challenges to dynamic stability, especially in the mediolateral direction [3].

The human body’s center of gravity (COG) is the imaginary point around which gravity appears to act. It is the point where the total mass of the body appears to be concentrated [4]. The COG is located 5 cm anterior to the second sacral vertebra and is displaced 5 cm horizontally and vertically during an average adult male step [5]. To prevent falls, keep your center of gravity above the support base, including your legs and assistive devices. For efficient gait both vertical and horizontal center of gravity movements should be minimized [5]. The balance could be stable, it can be unstable or dynamic [6]. Unchanged state of balance relies on proprioceptive data or input from; soles of the feet, spine (cervical), and sacroiliac joints [7]. To maintain postural balance information from sensorimotor system are combined along with input from eyes and inner ear.

Although, the foot size differs considerably across the population [8], yet given that humans are bipedal and have direct touch with the ground, feet may be crucial for maintaining balance throughout life [9]. There are either extremely few or no studies looking at the possibility that a person’s foot size might impact their balance. It was found in a study on children that greater balance was correlated with increased foot length [10].

Fatigue is expressed as a decrease in maximal muscle strength with prolonged exercise and is seen as decrease in physical performance [11]. When a person performs maximal exercise, a rapid decrease in muscle creatine phosphate and accumulation of metabolites such as lactate is observed [12,13] which affects the normal functioning of several brain structures [14,15]. The purpose of human movement is for the entire body to move forward through space, and the locomotion of individual segments is carried out in such a way as to maintain the vertical and lateral displacement of the COG to a minimum [16]. It has been documented that the gait cycle involves the exchange of kinetic and potential energy, which is an energy-efficient mode of movement [16, 17].

Sports training sessions represent a physiological stress that can cause temporary changes in physiological functions, metabolic parameters [18] and kinematic variables [19]. Therefore, fatigue is expected to result in biomechanical compensation and alter subjects’ normal movement patterns [20]. It further depends upon the type, duration, and intensity of training [21], and also personal factors such as medical conditions, age, and training level [19]. It is generally recognized that fatigue affects a variety of gait characteristics, including balance control [19, 22-24].

Normally instrumented walkways or multi cameras are used to analyze gait in strict laboratory conditions [25, 26]. Now a days to perform mobile gait test inertial measurement unit-based methods are being employed. A growing interest has been observed using IMU based data and algorithm for the analysis of gait, to be applied in the field of biomechanics, rehabilitation and sports science [27]. IMU based sensors are quite reliable, handy and cost effective to study and analyze the gait in both home and clinical settings [28]. Clinical validity and feasibility to measure spatiotemporal gait parameters of IMU based gait test has already been established in previous studies done on healthy young population and [29] Parkinson disease [30, 31].

Wearable sensors have the potential to be effectively utilized for analysis of gait in sports to analyze and improve the performance of athletes. Ambulatory gait techniques have been employed in previous studies to analyze segmental movements in golf [32], running [33], and baseball [34]. Wearable sensors have also been used in exercise coaching for the analysis of gait [35].

Wearable devices enable long-term continuous monitoring, paving the way for the development of accurate models for real-time fatigue monitoring. Hence, they are quite promising in monitoring the fatigue. Although there exist many methods to detect and monitor fatigue, there is still no absolute method to detect and monitor fatigue. In past many studies have been done on detection of fatigue in different sports using wearable technology, but there is hardly any study which have been done to detect fatigue in low intensity exercise or training. Therefore, it is important study the sensitivity of these sensor especially in low intensity exercise.

Therefore, we examined the sensitivity of wearable sensors in detection of fatigue induced by the low intensity football training in university football players. We collected data about somatic fatigue with wearable GPS tracking sensor, wearable heart rate monitoring sensor and self-reported RPE (Rate of Perceived Exertion) to confirm if the participant’s perceived exhaustions were connected to the walking dynamics.

## Methods

### Participants

For this study, twenty young, healthy male university football players were chosen as participants. None of the subjects had a history of any major muscle, neurological, cardiovascular, metabolic, or inflammatory condition, nor history of lower extremity trauma. Every participant practiced football twice a week and had normal or corrected-to-normal eyesight. Every participant completed a written informed consent form, and participation in the study was completely voluntary. This study was approved by the Institutional Review Board (IRB) at Seoul National University (SNU), Seoul, South Korea, protocol number 2402_001-025. The research protocol adhered to the ethical guidelines outlined by the IRB and was conducted in accordance with the principles of the Declaration of Helsinki. Informed consent was obtained from all participants prior to their participation in the study. The confidentiality and privacy of participants were strictly maintained throughout the research process. Any identifiable information was anonymized to ensure participant confidentiality.

### Wearable Sensor Gait Test

Data acquisition system and a Shoe-type IMU sensor-based gait analysis systems (DynaStab™, JEIOS, Korea) consisting of shoe-type data loggers (Smart Balance® SB-1, JEIOS, Korea) was used in the present study. An IMU sensor (IMU-3000TM, InvenSense, USA) integrated into a shoe-type data logger was capable of measuring tri-axial angular velocities (up to ±500° s− 1) and tri-axial acceleration (up to ±6 g) along three orthogonal axes [36, 37]. Outsoles of both shoes were fitted with IMU sensors and the collected data was transmitted via Bluetooth to the data acquisition system. Range of the shoe size was from 225 mm to 280 mm. The IMU sensors’ local coordinate system was set up with vertical, mediolateral, and anteroposterior orientations. (Fig. 1). IMU sensor was further used to collected data on spatiotemporal variables. All parameter of center of gravity of body was measured to get information about dynamic stability control.

**Fig 1.**
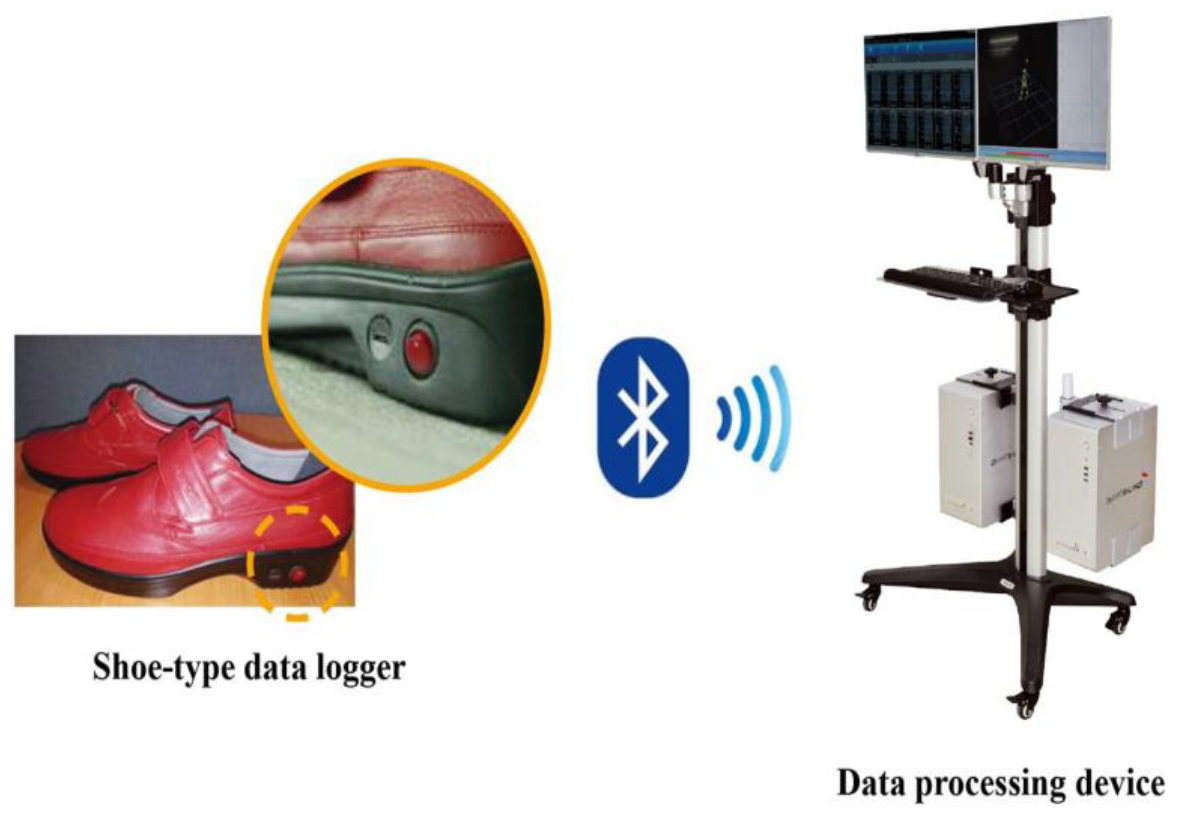
Shoe-type IMU Sensors Local Coordinate System

### Exercise Monitoring

Catapult vector, (Catapult, Australia) (Fig. 2), GPS players tracking device was used to monitor training parameters during football session. GPS (Global Positioning System) is widely accepted technology to asses competition and individual training workloads [38]. These GPS trackers are easy to use and does not need any additional installation or equipment [38], thus, have been used in research in across all range of sports. We used a customized harness and each device was deployed in the pocket of the harness. Devices were kept apart at a distance of 10 cm so that there is no interference occur between the signal of these devices. Polar heart rate monitors were used to measure exercise heart rate throughout the training sessions (Polar Electro Inc; www.polar.com/us-en) (Fig. 2).

**Fig 2.**
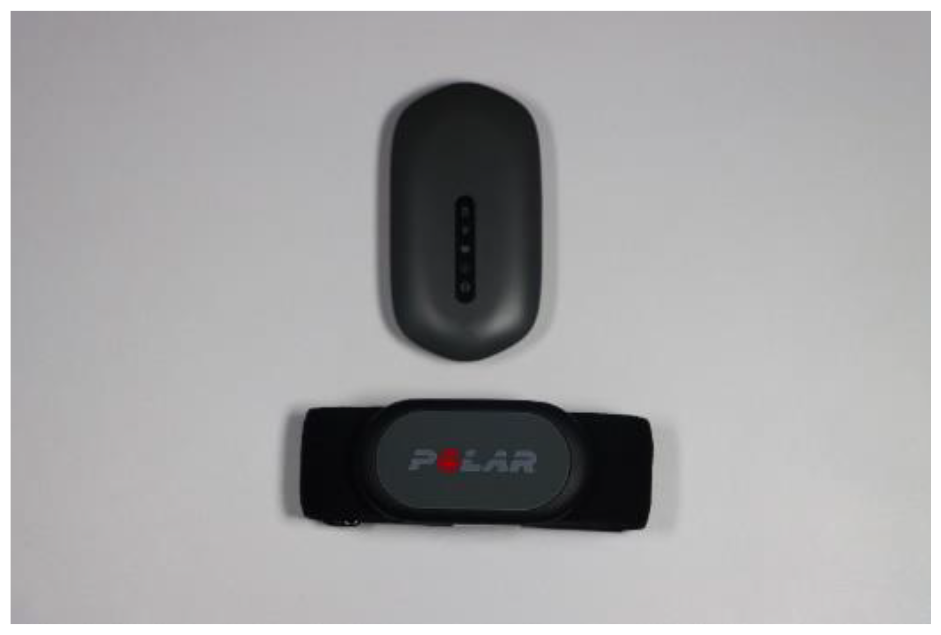
Wearable GSP Sensor Tracking (Catapult, Australia), Wearable Heart Rate Sensor (Polar, USA)

### Self-reported RPE

One way of measuring exhaustion and fatigue is self-perception. We used a rate of perceived exertion (RPE) scale to quantify fatigue. In this scale which was designed by Borgs, participants rate their level of exhaustion from scale of 1 (means no exertion) to 10 (means maximum exertion) [39]. Borgs scale is widely accepted to measure exhaustion and muscle fatigue after the football sessions [40, 41].

### Study Design

All twenty participants followed the experimental protocol which was divided in to three parts. In the first part pre-exercise measurements were recorded for all participants. Participants had to wear IMU sensors and walk on the treadmill at preferred moving speed without any interruption. Second part is the 90 minutes of football training sessions. In the third and last part participants have to repeat the gait test immediately after the end of football training sessions (Fig 3).

**Fig 3.**
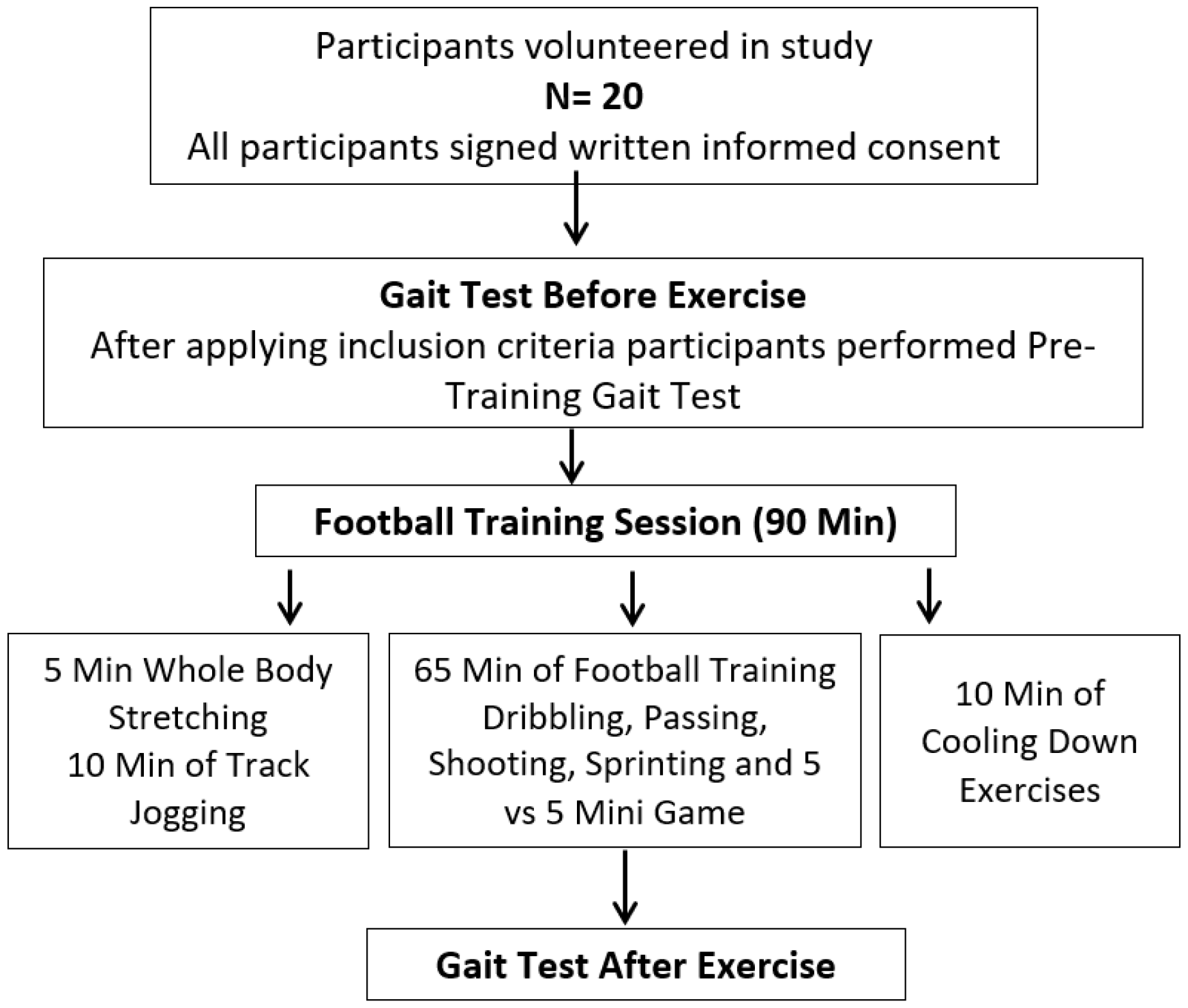
Experimental Protocol

### Fatigue Induced Low Intensity Football Training Sessions

Participants underwent two sessions of football training per week for a period 4 weeks. Drills involved complexity of movement involving shift in direction and speed. Training session were organized on regular football field for 90 minutes including 5 minutes whole body stretching, 10 minutes of track jogging, 65 minutes of main exercise which includes; Dribbling Drill, Passing Drill, Shooting Drill, Sprint Work Drill and 5 vs 5 mini game followed by cool-down exercise for 10 minutes. The volume of exercise and other exercise parameters were measured by GPS sensor, heart rate sensor and Borg scale of self-perceived exertion. (Table 1)

**Table 1.** Work Rate Tracing by GPS Sensor, Heart Rate Sensor and Self-Reported RPE.

|  | **Mean ± SD** |
| --- | --- |
| **Duration** | 90 min |
| **Total Distance (m)** | 6160.2 ± 2530.8 |
| **Max Heart rate (beat/min)** | 152.3 ± 63.1 |
| **Avg. Heart Rate (beat/min)** | 118.9 ± 47.8 |
| **Heart Rate (%Max)** | 75.5 ± 31.1 |
| **Borg RPE scale (1- 10)** | 4.3 ± 1.5 |

### Statistical Analysis

Collected data was analyzed and tested for normality. Descriptive data was presented as means and standard deviation. The level of significance was set at p ≤ 0.05). Since our data was not normal and sample size was small, we used non-parametric statistics. Wilcoxon signed rank test was used to analyze paired observations. Statistical analysis was processed with RStudio Team (2020). RStudio: Integrated Development for R. RStudio, PBC, Boston, MA URL http://www.rstudio.com/.

## Results

General and anthropometric data are reflected in (table 1 and 2). There was no withdrawal of any participants form the study. All football sessions were of low to moderate intensity as shown in table 1.

**Table 2.** Anthropometric Variables.

|  | **Mean ± SD** |
| --- | --- |
| **Age (years)** | 20.85±1.9 |
| **Height (cm)** | 176.16±4.39 |
| **Weight (kg)** | 69.97±6.02 |
| **BMI (kg/m<sup>2</sup>)** | 22.53±1.49 |
| **Shoes Size (mm)** | 268.53±6.76 |
| **Preferred walking speed (m/s)** | 3.19 |

### Spatio-Temporal Parameters Difference Between Before and After the Exercise

After analyzing the data, it has been observed that, after low to moderate intensity football training there has been significant change in most of the gait parameters except speed and stride length. During the analysis almost all spatio-temporal parameters were taken into consideration. Wilcoxson singed rank revealed that cadence was decreased (p < 0.0273) significantly post exercise. Fatigue induced exercise also led to decrease in the step speed but non-significantly. On the other hand, the stride length remained unchanged. Data further revealed that percentage of single support decreased significantly (p < 0.013), while percentage of double support increased significantly (p < 0.046). Significant increase in the time of toe off percentage (p<0.015) has also been observed along with significant increase (p<0.020) in gait asymmetry. Norm STL (normalized Stride length) remained unchanged (Table 3).

**Table 3.**
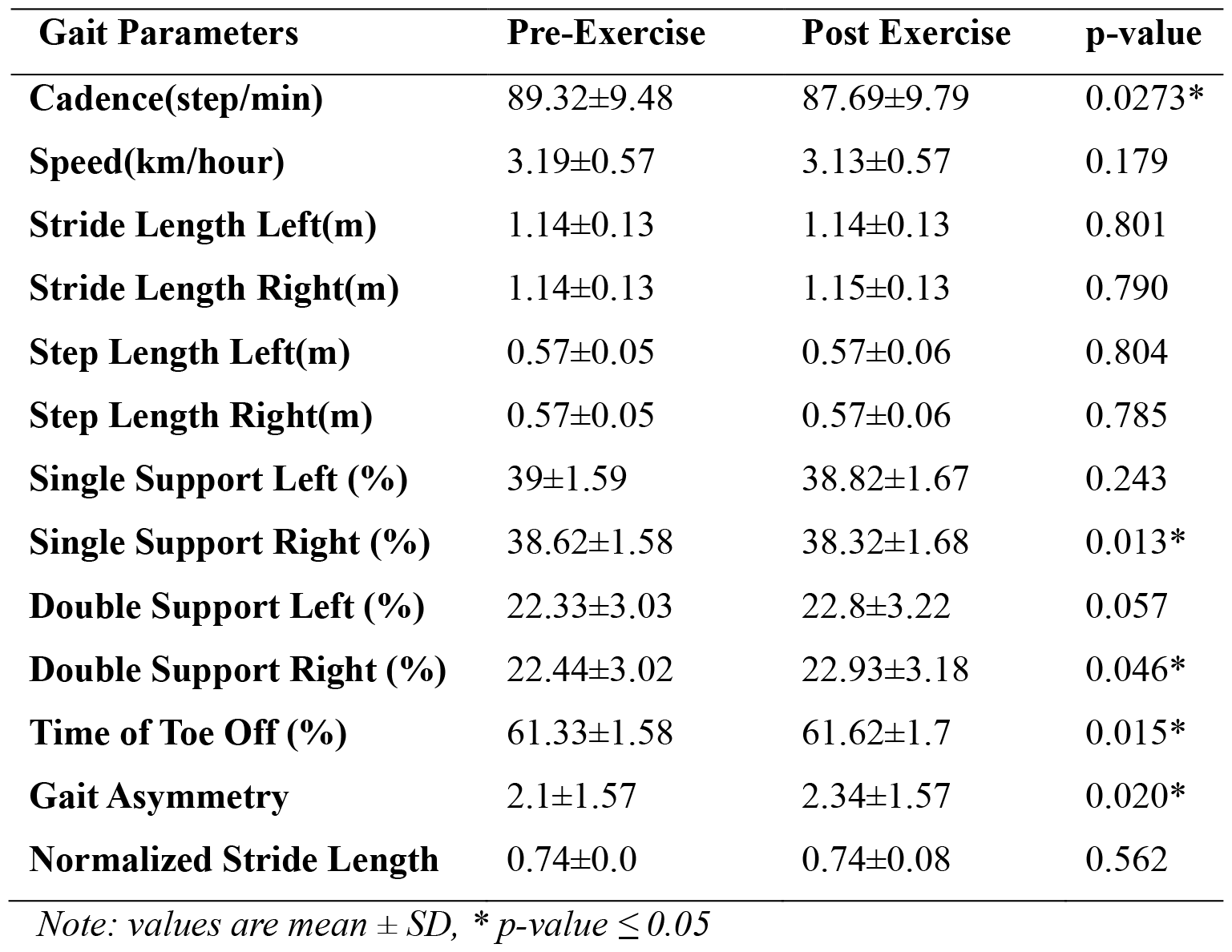
Spatio-temporal gait parameters pre and post exercise.

**Table 4.** Center of Gravity Parameters.

| COG Parameters | AX | Pre-Exercise | Post Exercise | p-value |
| --- | --- | --- | --- | --- |
| **Base of Support Width** |  | 0.05±0.03 | 0.05±0.03 | 0.688 |
| **Center of Gravity Distance (m)** | AP(X) | 0.1±0.02 | 0.1±0.02 | 0.278 |
|  | ML(Y) | 0.03±0.01 | 0.03±0.01 | 0.536 |
|  | SI(Z) | 0.03±0.01 | 0.03±0.01 | 0.083 |
| **Center of Gravity Velocity (m/s)** | AP(X) | 1.01±0.18 | 0.99±0.19 | 0.279 |
|  | ML(Y) | 0.33±0.1 | 0.34±0.1 | 0.533 |
|  | SI(Z) | 0.3±0.09 | 0.32±0.09 | 0.082 |
| **Center of Gravity Acceleration (m/s<sup>2</sup>)** | AP(X) | 0.3±0.09 | 0.32±0.09 | 0.082 |
|  | ML(Y) | 0.13±0.03 | 0.14±0.06 | 0.010* |
|  | SI(Z) | 0.24±0.06 | 0.24±0.06 | 0.250 |

*Note: values are mean ± SD, \*p-value ≤ 0.05, AP: anterior to posterior, ML: medial to lateral, SI: superior to inferior*

### Center of Gravity Parameters Difference Between Before and After Exercise

There were no significant differences on Center of Gravity (COG) parameters between pre-and post-gait test except central of gravity acceleration ML(Y) which increased significantly post exercise (p<0.01).

## Discussion

We examined the gait parameters of 20 participants following 90 min of low intensity football training. Gait characteristics calculated on fatigued trials were affected by large variance because effect of fatigue on gait parameters appeared to differ on amount of exhaustion achieved. Individual characteristics like age and degree of training might explain it. Our findings observed significant changes in cadence post exercise. Previous research has shown that fatigue affects motion behavior, which in turn affects kinematics [19, 20], [42, 43].

Rigorous exercise is linked to a marked decline in the capacity to maintain both the fluidity of walking and the bipodal upright posture [44]. In present study, even low to moderate intensity exercise affected the cadence, shown a decrease in the percentage of single support phase with consequent increase in percentage of double support associated with a decrease of time toe off and increase in gait asymmetry.

A previous study found that since it was difficult for the subjects to adjust to the ground and prepare for the push-off phase of gait, there may have been an increase in both stride length and step length to make up for the extended single support and terminal stance time [45]. On contrary to that, our findings reported only marginally increase in right stride length with no change in step length. Interestingly, we observed significant decrease in right single support duration indicating unhealthy gait. On the other hand, we have seen the significant increase in right double support duration indicating participants losing coordination and balance during dynamic gait post exercise. As stated in previous study, following the intrinsic foot core muscles exhaustion, an individual’s increase in stride length to compensate for the elongation in single support and terminal stance during the function of maintaining constant gait speed had a substantially bigger clinical impact [45]. However, in our study there was not much change in many gait parameters, possible because of low to moderate intensity of training, but we are able to detect significant changes in some of the critical parameter of gait test. Research has shown that stride length and step length are important variables that influence the likelihood of injuries, and that a shorter stride length is strongly associated with a higher risk of injuries [46].

There are potential benefits form the larger foot size on the human balance [47]. Since the center of gravity (COG) must be situated within the region of base of support (BOS) in order to sustain standing balance, prior research has demonstrated that the BOS is one of the primary elements impacting human balance while standing quietly, in addition to body weight. bigger feet often encompass bigger BOS areas, which increases COG movement flexibility and enhances balancing performance [48]. A greater base of support is necessary for those who are taller. Because of this, taller people typically have larger shoe sizes and longer feet. Previous studies done on foot size and height ratio have shown a high linear correlation between length of foot and height in both male and female [49, 50]. According to data from previous research, a bare footprint on a hard surface corresponds to 13.93% of a woman’s height and 14.35% of a man’s height [51]. Following a review of the literature on the subject, it was concluded that the foot length/stature ratio was consistently 15% throughout all populations [52,53].

Participants in our study can be considered to have normal foot length. We did not observe any change in BOS post exercise, in fact all parameter of COG remains unchanged except central of gravity acceleration ML(Y) which increased significantly post exercise (p<0.01). While foot size can play a role in determining the size and shape of the support surface for standing, it is not the determining factor in a person’s ability to balance, there are several other factors that can later balance and COG.

Greater propelling force is required for larger foot area and lower center of gravity [54], which results fatigue. Slow-twitch muscle fibers in humans are more resistant to fatigue, where as fast-twitch muscle fibers in humans may produce large amounts of power quickly [55]. The muscles’ capacity to generate force decreases both during and after a football training session. Anaerobic energy metabolism is crucial since physical activity is often associated with increased energy demands. As a result, this process causes an increase in blood lactate concentration and an increase in lactate generation in the working muscles. In addition, this led to a reduction in the force generated by muscular contractions, hence diminishing the muscle’s ability to sustain postural stability. The findings of previous study also demonstrated impaired postural control, which was often brought on by reduced proprioceptive or vestibular sensitivity and poorer muscle efficiency [56].

It is well established that a variety of factors, such as the buildup of metabolites inside muscle fibers or the insufficient motor command produced by the motor cortex, can contribute to fatigue. Muscle fatigue has no one cause and is very unique to the activity that is being done [57]. Contractile processes are responsible for producing skeletal muscular force, and malfunctions in any of the systems upstream of the cross-bridges can lead to onset of muscular fatigue, including the energy, vascular, ion, and nervous systems [58].

Muscle fatigue is specifically influenced by metabolic variables and fatigue reactants during contraction, such as heat shock protein (HSP), lactate, inorganic phosphate (Pi), reactive oxygen species (ROS), and hydrogen (H+) ions [59].

Exercise and fatigue not only cause ROS generation and ATP depletion, but they can also cause a localized or systemic inflammatory response. Leukocytes, IL-6, and TNF-α are promising biomarkers to assess inflammation in muscle fatigue [60].

Our study demonstrates that fatigue can be induced by even low to moderate exercise, within RPE range of 4.5 and at 60% of maximum heart rate. Fatigue induced by low to moderate-intensity exercise affects walking, and utilizing gait test through a wearable IMU sensor proves to be a highly sensitivity method for measuring accumulated fatigue in the human body, regardless of exercise intensity. Although determining the exact relationship between any of these characteristics and fatigue is challenging, their combination may be able to determine the presence of fatigue following exercise sessions.

Use of non-randomized trials hampered this investigation. Furthermore, only the gait of young, healthy males was examined. Future research should look at how fatigue affects women in order to determine any gender disparities. Lastly, as a further biomarker, we did not examine the muscle phosphocreatine readings. We advise doing a thorough gait study of these factors in future research and evaluating the impact of fatigue on other gait features.

## Conclusion

A portion of our hypotheses were partially confirmed. Variations in stride length and step width appear to be compensatory measures for reduced and compromised gait stability. The findings suggest that the continuous loss of muscular force production may be the cause of the modulation of gait. Furthermore, observed muscular exhaustion may indicate a higher chance of injury. Wearable IMU sensor gait test is considered a highly suitable method for measuring not only exercise-induced fatigue but also the physical deterioration resulting from accumulated fatigue during daily life. Fatigue is inevitable due to exhaustion form high intensity exercise and can be easily detected. It’s not that difficult to recognize when one feels fatigued, but understanding the physiological processes that lead to this state is a whole different story. In present study we can conclude that wearable IMU sensor gait test was found to be effective in detecting fatigue even in low to moderate intensity exercise. Nevertheless, in case of submaximal contraction (low to moderate intensity exercise) the onset of fatigue is most likely not connected with the termination of task. Since most of the daily tasks require submaximal contractions and are of low to moderate intensity, the onset of fatigue could not prevent an individual from completing a task (football training in present study), and wearable IMU sensor can detect fatigue in such activities using gait test.

## Data Availability

All data produced in the present study are available upon reasonable request to the authors

